# Concurrent changes in sleep and cognitive function during retirement transition: the Finnish Retirement and Aging Study

**DOI:** 10.1101/2024.05.14.24307322

**Authors:** Tea Teräs, Saana Myllyntausta, Jaana Pentti, Jesse Pasanen, Suvi Rovio, Sari Stenholm

**Affiliations:** Department of Public Health, University of Turku and Turku University Hospital, Turku, Finland; Centre for Population Health Research, University of Turku and Turku University Hospital; Turku, Finland; Department of Psychology and Speech-Language Pathology, University of Turku, Turku, Finland; Research Center of Applied and Preventive Cardiovascular Medicine, University of Turku, Turku, Finland; Clinicum, Faculty of Medicine, University of Helsinki, Finland; Research Services, Turku University Hospital and University of Turku, Finland

**Author notes:** Corresponding author: Tea Teräs, MD Department of Public Health, University of Turku, Turku, Finland.

**Keywords:** Cognitive function, Retirement, Sleep duration, Sleep difficulties, Accelerometry

## Abstract

**Background:** The transition to retirement has been shown to be accompanied by increased sleep duration and improved sleep quality. In addition, some studies suggest accelerated decline in cognitive function in post-retirement years. However, less is known about their interconnectedness. The aim of this study was to examine the concurrent changes in sleep and cognitive function during retirement transition.

**Methods:** The study population consisted of 250 public sector workers (mean age before retirement 63.1 years, standard deviation 1.4) from the Finnish Retirement and Aging study. The participants used a wrist-worn ActiGraph accelerometer, responded to the Jenkins Sleep Problem Scale and underwent cognitive testing annually before and after retirement. Computerized Cambridge Neuropsychological Test Automated Battery (CANTAB^®^) was used to evaluate learning and memory, working memory, sustained attention and information processing, executive function and cognitive flexibility and reaction time.

**Results:** Cognitive function improved in all cognitive domains, except for reaction time, during 1- year retirement transition period. The improvement was temporary in learning and memory, working memory and executive function and cognitive flexibility, which plateaued in post-retirement years. The participants were categorized into constantly short (49%), increasing (20%), decreasing (6%), and constantly mid-range (25%) sleep duration; and constantly without (36%), increasing (10%), decreasing (16%), and constantly with (38%) sleep difficulties. There were no associations between changes in sleep duration or sleep difficulties and cognitive function during retirement transition.

**Conclusions:** Cognitive function improves temporarily during transition to retirement, but the improvement is independent of changes in sleep characteristics.

---

Cognitive function changes during one’s lifespan. On average, cognitive function increases in the early years, stabilizes in middle-age, and starts to decline with advancing age.[1,2] The key cognitive changes with normal aging include decline in performance in tasks requiring rapid information processing for decision making, problem solving, and multitasking such as processing speed, working memory, and executive function.[3] On the other hand, so-called crystalized abilities such as vocabulary, verbal reasoning, and visual recognition are preserved well into old age.[3] With the aging population, the decrease of cognitive function becomes more apparent, and it would be valuable to find modifiable risk and protective factors affecting cognitive function trajectories with aging.

Retirement is a major life event in late middle-age resulting in numerous changes in everyday life. Retirement has been shown to associate with improved sleep [4–6] and psychological well-being [7], as well as to modify physical activity behavior.[8] However, the findings regarding changes in cognitive function are inconclusive. Some studies suggest that decline in cognitive function accelerates after retirement with a follow-up ranging from 4 to 18 years.[9–16] On the other hand, a recent meta-analysis showed that retirement does not associate with global cognitive function, but performance in memory-related tasks slightly decreases after retirement.[17] Several factors may explain these mixed findings. First, the length of follow-up as well as the characteristics of the study populations have varied markedly in terms of age and occupational background. Second, previous research has mainly focused on global cognitive function [13], a single cognitive domain [12,16], or only a few selected cognitive domains [10,11] rather than a wider spectrum of cognitive domains.

Moreover, it remains unclear how cognitive function changes during the transition to retirement, as previous research has mainly focused on trends of cognitive function before and after retirement rather than during the actual transition. Consequently, there is a need for further examination to identify subtle changes in different cognitive domains during the retirement transition years among participants from various occupations.

Previous studies have identified several factors that may modify the rate of cognitive decline after retirement including sex [10,11,18], occupation [12,14,19,20], job strain [21], retirement age [22–24], mental activities [13], and health.[25] However, the role of sleep on changing cognitive function during and after retirement is still ambiguous. We have previously shown that increasing and decreasing sleep difficulties are associated with a more pronounced decline in cognitive function during retirement transition during a time span of 5 years.[26] Apart from that, although it is known that sleep duration and difficulties associate with cognitive function,[27–30] the role of sleep on changes in cognitive function during the retirement transition has been scarcely examined.

This study aims to expand the previously studied association of retirement on cognitive function by narrowing the follow-up to annual examinations around retirement to study the short-term changes in multiple cognitive domains during the retirement transition. We also aim to examine whether changes in sleep duration and difficulties play a role in the change in cognitive function during retirement transition.

## METHODS

### Study population

The study population consisted of the participants of the Finnish Retirement and Aging Study (FIREA), an ongoing longitudinal study of older public sector employees in Finland established in 2013. Detailed description of the FIREA study design and implementation has been reported elsewhere.[31] Shortly, participants were first contacted 18 months prior to their estimated retirement date by sending them a questionnaire. Finnish-speaking participants living in Southwest Finland whose estimated retirement date was between 2017–2019, and who were still working were invited to participate in the clinical sub-study (n = 773). Of them, 290 participated in the first clinical examination. Thereafter, participants were followed up with annual measurements including questionnaires and clinical and accelerometer measurements. To determine the timing of retirement, working status was inquired during an annual clinical visit. Data was then centered around the retirement transition and presented as pre-retirement (waves -2 to -1), retirement transition (waves - 1 to 1) and post-retirement (waves 1 to 2) periods. For this study, the participant had to have information on cognitive function and sleep characteristics from study waves before and after retirement (i.e., waves -1 and 1). This resulted in a study sample of 250 participants.

Informed consent was obtained from all participants. The FIREA study was conducted in accordance with the Helsinki declaration and was approved by the Ethics Committee of Hospital District of Southwest Finland.

### Cognitive function

Cognitive function was evaluated annually during clinical examinations utilizing computerized Cambridge Neuropsychological Test Automated Battery (CANTAB^®^; https://www.cambridgecognition.com/cantab/). CANTAB is a widely used standardized test battery which comprises of a variety of different cognitive tests covering multiple cognitive domains including working memory; executive function; learning; visual, verbal, and episodic memory; attention, information processing and reaction time; social and emotion recognition, decision making, and response control. The tests used in this study were Paired Associates Learning (PAL) for *visual memory and associative learning* (hereafter *learning and memory*), Spatial Working Memory (SWM) for *working memory*, Rapid Visual Information Processing (RVP) for *sustained attention and information processing*, Attention Switching Task (AST) for *executive function and cognitive flexibility* and Reaction Time (RTI) for *reaction time*. More thorough description of the tests can be found elsewhere.[32]

Each CANTAB^®^ subtest produces several outcome variables. For data reduction and to gain summary variables that would explain most of the variation within the data set, we created Z-score based summary scores for each CANTAB subtest. First, each variable was standardized and converted so that a higher value reflects better cognitive function. After that, subtest specific variables were summed and divided by the number of variables from that specific subtest. At baseline, each individual variable was transformed into a scale with a mean of 0 and a standard deviation (SD) of 1. In the follow-up measurements, the standardization was conducted in respect to the baseline distribution (mean and standard deviation) and these values were used to create the summary scores for specific subtests.

### Sleep characteristics

Sleep duration was evaluated objectively with accelerometry. The wrist-worn triaxial wActiSleep- BT accelerometer by ActiGraph (Pensacola, Florida, US) was initialized to record movements during sleep and wakefulness at 80 Hz. The participants received the device via mail before the cognitive testing and were instructed to wear it continuously on their non-dominant wrist for 24- hours per day for at least seven days and nights (including at least two workdays and two free days while still working). The data handling and checking procedures as well as the used algorithms have been described in detail elsewhere.[4] Sleep duration was categorized into short (<7 hours), average (7-<9 hours), and long (≥9 hours) sleep duration. As there were only two (0.87%) long sleepers before retirement and none after, long-sleepers were excluded from the analyzes regarding sleep duration. Based on participant’s sleep duration immediately before retirement (study wave -1) and after retirement (study wave +1), they were then further categorized into 1) constantly short sleep duration, 2) increasing sleep duration (i.e. short sleepers before retirement and average sleepers after), 3) decreasing sleep duration (i.e. average sleepers before retirement and short sleepers after) and 4) constantly average sleep duration.

Sleep difficulties were evaluated with Jenkins Sleep Problem Scale, a four-item survey including questions about difficulties in falling asleep, difficulties in maintaining sleep during the night, waking up too early in the morning, and nonrestorative sleep.[33] The response categories for each item were 1) never, 2) 1-3 nights per month, 3) 1 night per week, 4) 2–4 nights per week, 5) 5–6 nights per week, and 6) nearly every night. Items of Jenkins Sleep Problem Scale correspond to the Diagnostic and Statistical Manual of Mental Disorders (DSM) 5 diagnostic criteria for insomnia (excluding nonrestorative sleep) [34]. As the DSM-5 defines insomnia as any of these symptoms occurring at least three nights per week, we similarly defined sleep difficulty as any of the items occurring at least 2–4 night per week. Based on the responses immediately before (study wave -1) and after retirement (study wave +1), the participants were then categorized into 1) constantly without sleep difficulties, 2) increasing sleep difficulties (i.e. no difficulties before retirement but difficulties after), 3) decreasing sleep difficulties (i.e. difficulties before retirement but none after), and 4) constantly with sleep difficulties.

### Covariates

The covariates used in this study were age, sex, occupational position, depression, job strain, and alcohol risk use. These covariates were selected based on their known association with sleep characteristics and cognitive function. Covariates were evaluated at the study phase immediately before retirement (study wave -1).

Participants’ date of birth, sex, and occupational status were obtained from the pension insurance institute for the municipal sector in Finland. Occupational status was categorized based on the International Standard Classification of Occupations (ISCO) into three groups according to the occupational titles: managers and professionals as administrative (ISCO classes 1-2), associate professionals and office workers as professional / executive (ISCO classes 3-4), and service and manual workers as clerical / support (ISCO classes 5-9).

Depression, job strain, and alcohol risk use were obtained from the questionnaires. Depression was evaluated with Beck Depression Inventory [35] with a cut-off point of 10 or more indicating mild or severe depression. Job strain was measured using scales of job control and job demands from the shorter version of the Job Content Questionnaire [36,37] using the median values from the entire

FIREA cohort as the cutoff points (job control 3.75 and job demands 3.2) to identify the participants with job strain (a high “demands” and a low “control” score). Alcohol risk use was defined as >16 drinks/week for women and >24 drinks/week for men corresponding with the lower limit for heavy use of alcohol set by the Finnish Ministry of Health and Social Affairs.[38]

### Statistical analyses

Sample characteristics before retirement are shown as percentages for categorical variables and means and standard deviations (SD) for continuous variables.

To estimate the mean level (95% confidence limits) of each cognitive domain in pre-retirement years (study waves -2 and -1) and post-retirement years (study waves +1 and +2), we performed linear regression analyses with generalized estimating equations (GEE) with an exchangeable correlation structure. The GEE model controls for intraindividual correlation between repeated measures and assumes that measurements are missing completely at random.[39,40]

Mean levels of each cognitive domain by sleep duration and sleep difficulty status before retirement (wave -1) were examined with analysis of variance. The association between changes in sleep duration and difficulties and changes in cognitive function during retirement transition (waves -1 to 1) were examined with GEE models, which included a “sleep group × time (i.e. cognitive function before or after retirement)” interaction term for group comparisons. The analyses were initially adjusted for age, sex, and occupational position. The analyzes were further adjusted for depression, job strain, and alcohol risk use.

All statistical analyses were performed using SAS version 9.4 (SAS Institute Inc., Cary, NC, USA).

## RESULTS

The pre-retirement (study wave -1) characteristics of the study population are shown in **Table 1**. The mean age of the participants at the study phase before retirement was 63.1 years (SD 1.1) and the majority of participants were woman (84%). Each occupational group was represented rather equally: 35% of the participants were in administrative, 34% in professional / executive, and 32% in the clerical / support occupations. Impaired sleep was relatively common before retirement: 53% of the participants reported sleep difficulties and 68% were short sleepers.

**Table 1.** Characteristics of the study population before retirement (N=250)

|  | Mean | SD |
| --- | --- | --- |
| Age | 63.1 | 1.1 |
|  | n | % |
| Sex |  |  |
| Male | 40 | 16 |
| Female | 210 | 84 |
| Occupational position |  |  |
| Administrative | 87 | 35 |
| Professional / executive | 84 | 34 |
| Clerical / support | 79 | 32 |
| Depression | 28 | 11 |
| Job strain | 43 | 19 |
| Alcohol risk use | 25 | 10 |
| Sleep difficulties | 131 | 53 |
| Sleep duration |  |  |
| <7 hours | 162 | 68 |
| 7-<9 hours | 74 | 31 |
| ≥9 hours | 2 | 0.8 |

The changes in each cognitive domain before, during and after retirement transition are shown in **Figure 1**. In terms of *learning and memory*, there was a clear improvement during retirement transition (p=0.003), which plateaued after retirement. Similarly, *working memory* improved during retirement transition (p=0.014), but remained stable during first post-retirement years. *Sustained attention and information processing* as well as *executive function and cognitive flexibility* improved throughout the follow-up period with an accelerated improvement during retirement transition (p=0.012 and p=0.015, respectively). *Reaction time* stayed at the same level before and after retirement and no improvement was observed during retirement transition (p=0.51). When further adjusted for depression, job strain, and alcohol consumption the results remained similar (**Supplemental Figure 1**).

**Figure 1.**
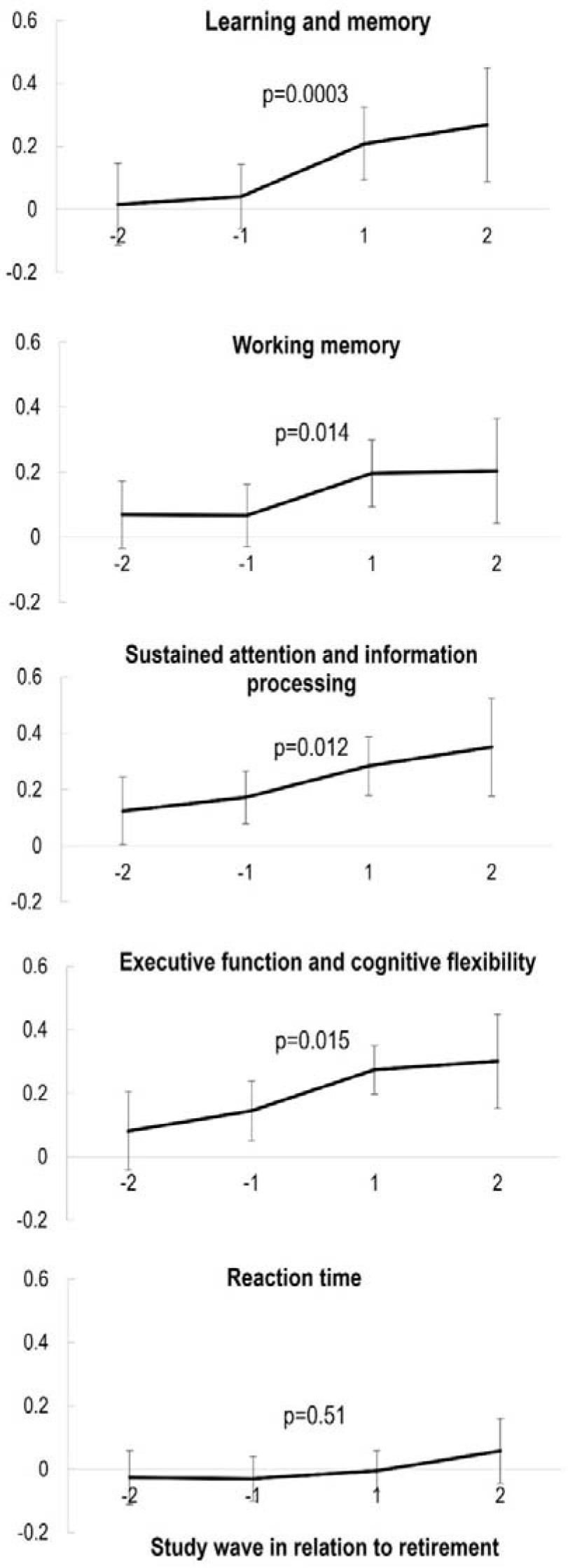
Mean level and their 95% confidence intervals in each cognitive domain before and after retirement. Adjusted for age, sex, and occupational position. The p-values that are shown describe the change in cognitive function during the retirement transition (wave -1 to wave +1).

**Table 2** shows the pre-retirement association of sleep and cognitive domains. Those with mid-range sleep had better performance in *executive function and cognitive flexibility* than short sleepers (p=0.014). There was also a tendency towards better *working memory* and *sustained attention and information processing* among mid-range sleepers compared to short sleepers, but these differences did not reach statistical significance level 0.05. On the contrary, those with short sleep duration had better *learning and memory* than mid-range sleepers (p=0.044). There were no differences in the level of cognitive domains between those with and without sleep difficulties before retirement.

**Table 2.** Mean levels of each cognitive domain by sleep duration and sleep difficulty status before retirement.

|  | Mean | 95% CI |  | p value |
| --- | --- | --- | --- | --- |
| **Learning and memory** |  |  |  |  |
| Short sleep duration | 0.11 | -0.02 | 0.23 | 0.044 |
| Mid-range sleep duration | -0.08 | -0.26 | 0.09 |  |
| No sleep difficulties | 0.08 | -0.06 | 0.21 | 0.469 |
| Sleep difficulties | 0.02 | -0.13 | 0.16 |  |
| **Working memory** |  |  |  |  |
| Short sleep duration | 0.07 | -0.05 | 0.18 | 0.092 |
| Mid-range sleep duration | 0.21 | 0.06 | 0.36 |  |
| No sleep difficulties | 0.13 | 0.01 | 0.25 | 0.504 |
| Sleep difficulties | 0.07 | -0.07 | 0.21 |  |
| **Sustained attention and information processing** |  |  |  |  |
| Short sleep duration | 0.15 | 0.04 | 0.26 | 0.059 |
| Mid-range sleep duration | 0.31 | 0.16 | 0.47 |  |
| No sleep difficulties | 0.18 | 0.05 | 0.31 | 0.812 |
| Sleep difficulties | 0.20 | 0.08 | 0.31 |  |
| **Executive function and cognitive flexibility** |  |  |  |  |
| Short sleep duration | 0.11 | 0.01 | 0.21 | 0.014 |
| Mid-range sleep duration | 0.30 | 0.19 | 0.41 |  |
| No sleep difficulties | 0.15 | 0.03 | 0.27 | 0.966 |
| Sleep difficulties | 0.15 | 0.03 | 0.28 |  |
| **Reaction time** |  |  |  |  |
| Short sleep duration | -0.06 | -0.17 | 0.04 | 0.512 |
| Mid-range sleep duration | -0.02 | -0.14 | 0.10 |  |
| No sleep difficulties | -0.07 | -0.18 | 0.03 | 0.527 |
| Sleep difficulties | -0.03 | -0.15 | 0.08 |  |

*Notes:* Adjusted for age, sex, and occupational position. Group sizes are: short sleep duration n=157, mid-range sleep duration n=71, no sleep difficulties n=115, and sleep difficulties n=127.

In terms of changes in sleep during retirement transition, 49% had constantly short sleep duration, 19% increased, 7% decreased, and 25% had constantly mid-range sleep duration. In addition, 36% remained without sleep difficulties, 10 % increased, 16% decreased, and 38% had constantly sleep difficulties. Age, sex, and occupational position adjusted results for changes in the studied cognitive domains during retirement transition within the sleep duration change groups are shown in **Table 3**. No statistically significant differences between the sleep duration change groups were observed.

**Table 3.**
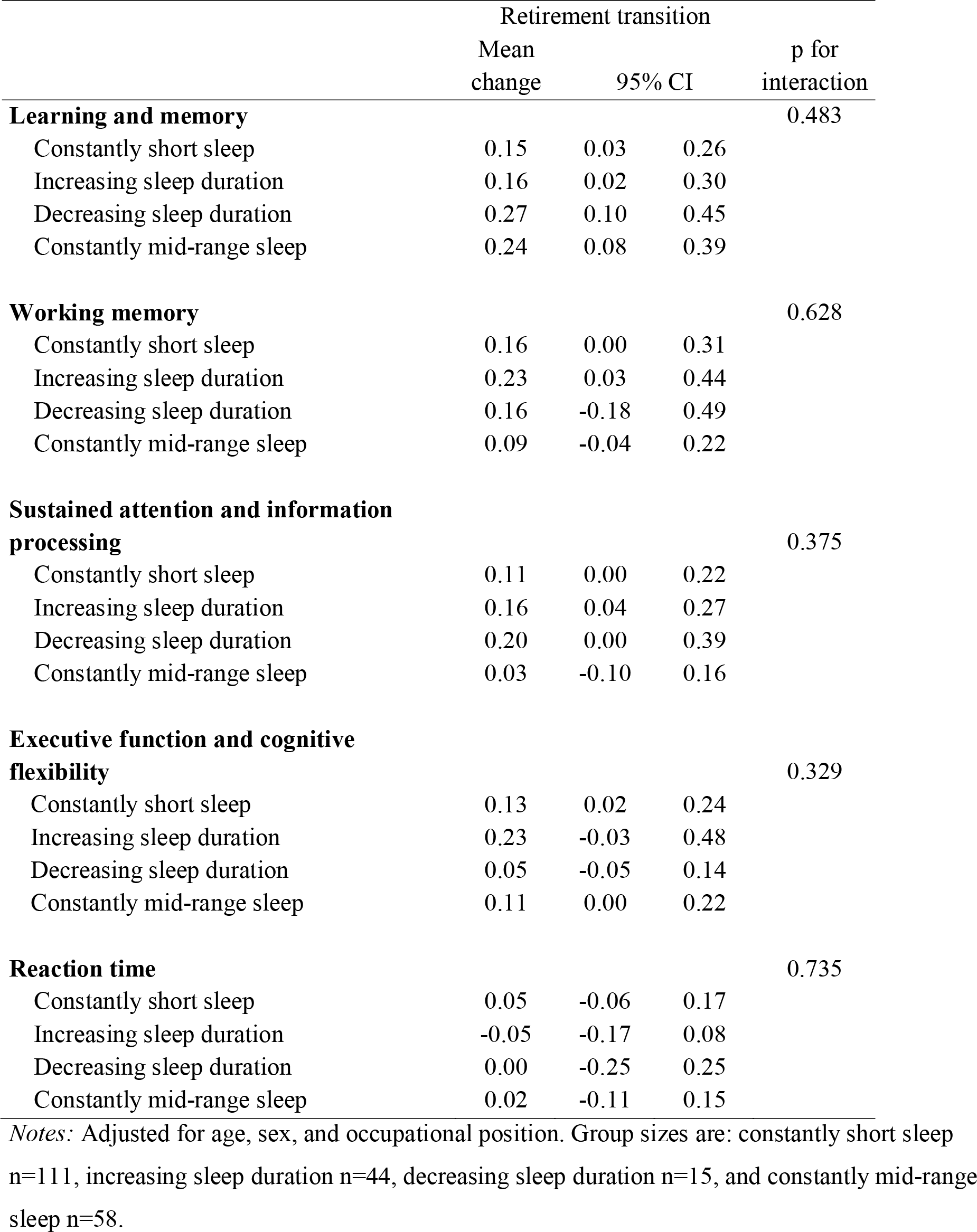
Mean change in each cognitive domain during retirement transition by sleep duration change groups.

|  | Retirement transition |  |  |  |
| --- | --- | --- | --- | --- |
|  | Mean<br>change | 95% CI |  | p for<br>interaction |
| **Learning and memory** |  |  |  | 0.483 |
| Constantly short sleep | 0.15 | 0.03 | 0.26 |  |
| Increasing sleep duration | 0.16 | 0.02 | 0.30 |  |
| Decreasing sleep duration | 0.27 | 0.10 | 0.45 |  |
| Constantly mid-range sleep | 0.24 | 0.08 | 0.39 |  |
| **Working memory** |  |  |  | 0.628 |
| Constantly short sleep | 0.16 | 0.00 | 0.31 |  |
| Increasing sleep duration | 0.23 | 0.03 | 0.44 |  |
| Decreasing sleep duration | 0.16 | -0.18 | 0.49 |  |
| Constantly mid-range sleep | 0.09 | -0.04 | 0.22 |  |
| **Sustained attention and information processing** |  |  |  | 0.375 |
| Constantly short sleep | 0.11 | 0.00 | 0.22 |  |
| Increasing sleep duration | 0.16 | 0.04 | 0.27 |  |
| Decreasing sleep duration | 0.20 | 0.00 | 0.39 |  |
| Constantly mid-range sleep | 0.03 | -0.10 | 0.16 |  |
| **Executive function and cognitive flexibility** |  |  |  | 0.329 |
| Constantly short sleep | 0.13 | 0.02 | 0.24 |  |
| Increasing sleep duration | 0.23 | -0.03 | 0.48 |  |
| Decreasing sleep duration | 0.05 | -0.05 | 0.14 |  |
| Constantly mid-range sleep | 0.11 | 0.00 | 0.22 |  |
| **Reaction time** |  |  |  | 0.735 |
| Constantly short sleep | 0.05 | -0.06 | 0.17 |  |
| Increasing sleep duration | -0.05 | -0.17 | 0.08 |  |
| Decreasing sleep duration | 0.00 | -0.25 | 0.25 |  |
| Constantly mid-range sleep | 0.02 | -0.11 | 0.15 |  |

Notes: Adjusted for age, sex, and occupational position. Group sizes are: constantly short sleep n=111, increasing sleep duration n=44, decreasing sleep duration n=15, and constantly mid-range sleep n=58.

The results remained similar when further adjusted for depression, job strain and alcohol consumption. Similarly, **Table 4** shows the age, sex, and occupational position adjusted results for changes in the cognitive domains during retirement transition in the sleep difficulty change groups. Again, no statistically significant differences were found between the groups, and the further adjustments for depression, job strain and alcohol risk use did not change the results.

**Table 4.** Mean change in each cognitive domain during retirement transition by sleep difficulty change groups.

|  | Retirement transition |  |  |  |
| --- | --- | --- | --- | --- |
|  | Mean<br>change | 95% CI |  | p for<br>interaction |
| **Learning and memory** |  |  |  | 0.574 |
| Constantly without sleep difficulties | 0.15 | 0.02 | 0.27 |  |
| Increasing sleep difficulties | 0.06 | -0.13 | 0.25 |  |
| Decreasing sleep difficulties | 0.18 | -0.01 | 0.36 |  |
| Constantly with sleep difficulties | 0.21 | 0.09 | 0.32 |  |
| **Working memory** |  |  |  | 0.420 |
| Constantly without sleep difficulties | 0.14 | -0.01 | 0.29 |  |
| Increasing sleep difficulties | 0.10 | -0.05 | 0.26 |  |
| Decreasing sleep difficulties | -0.01 | -0.23 | 0.21 |  |
| Constantly with sleep difficulties | 0.19 | 0.05 | 0.32 |  |
| **Sustained attention and information**<br>**processing** |  |  |  | 0.346 |
| Constantly without sleep difficulties | 0.17 | 0.04 | 0.29 |  |
| Increasing sleep difficulties | 0.16 | -0.06 | 0.38 |  |
| Decreasing sleep difficulties | 0.07 | -0.07 | 0.22 |  |
| Constantly with sleep difficulties | 0.05 | -0.06 | 0.16 |  |
| **Executive function and cognitive**<br>**flexibility** |  |  |  | 0.523 |
| Constantly without sleep difficulties | 0.14 | 0.01 | 0.27 |  |
| Increasing sleep difficulties | 0.03 | -0.12 | 0.18 |  |
| Decreasing sleep difficulties | 0.24 | -0.04 | 0.52 |  |
| Constantly with sleep difficulties | 0.12 | 0.02 | 0.21 |  |
| **Reaction time** |  |  |  | 0.299 |
| Constantly without sleep difficulties | 0.08 | -0.03 | 0.19 |  |
| Increasing sleep difficulties | -0.02 | -0.16 | 0.11 |  |
| Decreasing sleep difficulties | 0.11 | -0.04 | 0.26 |  |
| Constantly with sleep difficulties | -0.03 | -0.16 | 0.09 |  |

Notes: Adjusted for age, sex, and occupational position. Group sizes are: constantly without sleep difficulties n=88, increasing sleep difficulties n=23, decreasing sleep difficulties n=39, and constantly sleep difficulties n=92.

## DISCUSSION

In this study of retiring workers, we studied how cognitive function changes during retirement transition by addressing various cognitive domains. Additionally, we studied whether changes in sleep duration and difficulties are associated with the changes in different cognitive function domains. By utilizing annually collected data on cognitive function, we showed that cognitive function improves during the transition to retirement. Of the studied cognitive domains, this transient improvement was evident in learning and memory as well as in working memory. In addition, sustained attention and information processing as well as executive function and cognitive flexibility showed improvement throughout the follow-up from pre-retirement years to first post-retirement years. No change was observed in reaction time. When studying the role of sleep in the changes in cognitive function, we found that changes in sleep duration or sleep difficulties were not associated with the observed improvements in cognitive function.

The improved cognitive function during retirement transition is an interesting finding, as it would suggest that something hinders the employees from reaching their full cognitive potential whilst still in employment. The finding is in line with one previous study,[23] although most previous studies have found no such effect. The discrepancy in our and previous findings could be due to the different time frame of cognitive measurements as in most studies measurements have been biennial or even with longer follow-ups as their focus has been more on post-retirement years.[9–13,15,18–22,24,25] Celidoni et al. showed in a multi-national European study sample of over 50-year-olds that statutory retirement is at first beneficial to cognitive function, but it starts to decline in post-retirement years.[23] Similarly, Bonsang et al. showed that the negative effect of retirement on cognitive function is not instantaneous in a study sample of over 50-year olds living in the United States, but found no improvement during retirement transition.[16] Our results are in line with Celidoni et al. and Bonsang et al. but, they both focused only on memory function. The added value of our study is that we were able to study changes in multiple cognitive function domains including learning and memory, working memory, sustained attention and information processing, as well as executive function and cognitive flexibility, and reaction time, and found that performance in all studied cognitive domains, apart from reaction time, improved during retirement transition.

In the current study, we also examined whether sleep duration or sleep difficulties would be associated with short-term changes in cognitive function during retirement transition, since sleep duration has shown to increase and sleep difficulties to decrease during retirement transition [4,9,41,42] Moreover, only few previous studies have linked sleep duration[29,30,43] and sleep difficulties[29,32] to changes in cognitive function. However, we did not find any differences between the sleep duration or difficulty groups in terms of changes in cognitive function. This was somewhat unexpected, as we have previously shown, by using the long-term Whitehall II study, that increasing and decreasing sleep difficulties are associated with accelerated decline in cognitive function during retirement transition.[26] However, in that study the cognitive function measurements were 5 years apart and we observed cognitive function to decline only after post-retirement retirement years. Additionally, cognitive function was only evaluated in relation to inductive reasoning and verbal memory rather than a wider range of cognitive domains as was done in this study. The findings of the current study suggest that other factors than sleep characteristics explain the observed change in cognitive function during retirement transition, and this warrants further examination.

This study has several strengths. With annual examinations, we were able to focus more specifically on the transition to retirement unlike most previous studies. Cognitive function was measured with computerized standardized test battery, which can detect even small changes in cognitive function, and allowed us to study multiple individual cognitive domains. Additionally, we were able to utilize information on accelerometer-measured sleep duration, which provides more accurate estimation of one’s sleep duration.

There are also some limitations that need to be addressed. As the cognitive function measurement were repeated annually, some practice effect might have occurred, which has also been observed earlier with CANTAB test battery [44] However, many commonly used pen-and-paper cognitive neuropsychological tests show similar test-retest reliability.[45] Furthermore, compared to the traditional non-computerized test batteries, computerized batteries are also more precise in recording for example latency times and accuracy. Simultaneously, they can be more easily optimized so that factors such as ceiling effect does not violate the data, and thus, computerized tests are specifically suitable for relatively cognitively healthy cohorts like ours. Finally, given that the FIREA clinical study population was healthier and had higher occupational status compared to the FIREA survey-only participants [31] and they were able to work until moving to statutory retirement, the generalizability of the findings to aging workers in general may be limited.

## Conclusions

We found cognitive function to improve during the retirement transition. This improvement was independent from changes in sleep duration and sleep difficulties. This finding suggests that employees have unused cognitive reserve, and future studies should focus on determinants behind the now observed changes in cognitive function. Identifying targetable determinants behind this change might allow employees to capitalize the unused cognitive reserve when still in working life and therefore improve their productivity at work or even prolong their work-lives.

## Acknowledgements

The authors want to thank the FIREA participants for their willingness to participate in the study and the FIREA study staff members for their contribution in the data collection.

## Ethical Approval

The FIREA study was conducted in accordance with the Helsinki declaration and was approved by the Ethics Committee of Hospital District of Southwest Finland (ETMK: 84/1801/2014).

## Data Availability

Anonymized partial datasets of the FIREA study are available by application with bona fide researchers with an established scientific record and bona fide organizations. In case of data requests, please contact the principal investigator Sari Stenholm.

## REFERENCES

[1] Singh-Manoux A, Kivimaki M, Glymour MM, Elbaz A, Berr C, Ebmeier KP, et al. Timing of onset of cognitive decline: results from Whitehall II prospective cohort study. BMJ 2012;344:d7622. 10.1136/bmj.d7622 [doi].

[2] Van Der Willik KD, Licher S, Vinke EJ, Knol MJ, Darweesh SKL, Van Der Geest JN, et al. Trajectories of Cognitive and Motor Function Between Ages 45 and 90 Years: A Population-Based Study. J Gerontol A Biol Sci Med Sci 2021;76:297. 10.1093/GERONA/GLAA187.

[3] Murman DL. The Impact of Age on Cognition. Semin Hear 2015;36:111–21. 10.1055/S-0035-1555115.

[4] Myllyntausta S, Pulakka A, Salo P, Kronholm E, Pentti J, Vahtera J, et al. Changes in Accelerometer-Measured Sleep during the Transition to Retirement: The Finnish Retirement and Aging (FIREA) study. Sleep 2020. zsz318 [pii].

[5] Hagen EW, Barnet JH, Hale L, Peppard PE. Changes in Sleep Duration and Sleep Timing Associated with Retirement Transitions. Sleep 2016;39:665–73. 10.5665/SLEEP.5548.

[6] Myllyntausta S, Salo P, Kronholm E, Pentti J, Kivimaki M, Vahtera J, et al. Changes in Sleep Difficulties During the Transition to Statutory Retirement. Sleep 2018;41:10.1093/sleep/zsx182. 10.1093/sleep/zsx182 [doi].

[7] Lahdenperä M, Virtanen M, Myllyntausta S, Pentti J, Vahtera J, Stenholm S. Psychological Distress During the Retirement Transition and the Role of Psychosocial Working Conditions and Social Living Environment. J Gerontol B Psychol Sci Soc Sci 2022;77:135–48. 10.1093/GERONB/GBAB054.

[8] Suorsa K, Leskinen T, Pasanen J, Pulakka A, Myllyntausta S, Pentti J, et al. Changes in the 24-h movement behaviors during the transition to retirement: compositional data analysis. Int J Behav Nutr Phys Act 2022;19. 10.1186/S12966-022-01364-3.

[9] Xue B, Cadar D, Fleischmann M, Stansfeld S, Carr E, Kivimaki M, et al. Effect of retirement on cognitive function: the Whitehall II cohort study. Eur J Epidemiol 2018;33:989–1001. 10.1007/s10654-017-0347-7 [doi].

[10] Oi K. Does Gender Differentiate the Effects of Retirement on Cognitive Health? Res Aging 2019;41:575–601. 10.1177/0164027519828062/ASSET/IMAGES/LARGE/10.1177_0164027519828062-FIG3.JPEG.

[11] Hamm JM, Heckhausen J, Shane J, Lachman ME. Risk of Cognitive Declines With Retirement: Who Declines and Why? Psychol Aging 2020;35:449. 10.1037/PAG0000453.

[12] Carr DC, Willis R, Kail BL, Carstensen LL. Alternative Retirement Paths and Cognitive Performance: Exploring the Role of Preretirement Job Complexity. Gerontologist 2020;60:460. 10.1093/GERONT/GNZ079.

[13] Lee Y, Chi I, Palinkas LA. Retirement, Leisure Activity Engagement, and Cognition Among Older Adults in the United States. J Aging Health 2019;31:1212–34. 10.1177/0898264318767030/ASSET/IMAGES/LARGE/10.1177_0898264318767030-FIG1.JPEG.

[14] Meng A, Nexø MA, Borg V. The impact of retirement on age related cognitive decline – a systematic review. BMC Geriatr 2017;17. 10.1186/S12877-017-0556-7.

[15] De Grip A, Dupuy A, Jolles J, Van Boxtel M. Retirement and cognitive development in the Netherlands: Are the retired really inactive? Econ Hum Biol 2015;19:157–69. 10.1016/J.EHB.2015.08.004.

[16] Bonsang E, Adam S, Perelman S. Does retirement affect cognitive functioning? J Health Econ 2012;31:490–501. 10.1016/J.JHEALECO.2012.03.005.

[17] Alvarez-Bueno C, Cavero-Redondo I, Jimenez-Lopez E, Visier-Alfonso ME, Sequi- Dominguez I, Martinez-Vizcaino V. Effect of retirement on cognitive function: a systematic review and meta-analysis. Occup Environ Med 2021;78:761–8. 10.1136/OEMED-2020-106892.

[18] Atalay K, Barrett GF, Staneva A. The effect of retirement on elderly cognitive functioning. J Health Econ 2019;66:37–53. 10.1016/J.JHEALECO.2019.04.006.

[19] Fisher GG, Infurna FJ, Grosch J, Stachowski A, Faul JD, Tetrick LE. Mental Work Demands, Retirement, and Longitudinal Trajectories of Cognitive Functioning. J Occup Health Psychol 2014;19:231. 10.1037/A0035724.

[20] Finkel D, Andel R, Gatz M, Pedersen NL. The Role of Occupational Complexity in Trajectories of Cognitive Aging Before and After Retirement. Psychol Aging 2009;24:563. 10.1037/A0015511.

[21] Nilsen C, Nelson ME, Andel R, Crowe M, Finkel D, Pedersen NL. Job Strain and Trajectories of Cognitive Change Before and After Retirement. J Gerontol B Psychol Sci Soc Sci 2021;76:1313–22. 10.1093/GERONB/GBAB033.

[22] Hale JM, Bijlsma MJ, Lorenti A. Does postponing retirement affect cognitive function? A counterfactual experiment to disentangle life course risk factors. SSM Popul Health 2021;15. 10.1016/J.SSMPH.2021.100855.

[23] Celidoni M, Dal Bianco C, Weber G. Retirement and cognitive decline. A longitudinal analysis using SHARE data. J Health Econ 2017;56:113–25. 10.1016/J.JHEALECO.2017.09.003.

[24] Grotz C, Meillon C, Amieva H, Stern Y, Dartigues JF, Adam S, et al. Why is later age at retirement beneficial for cognition? Results from a French population-based study. Journal of Nutrition, Health and Aging 2016;20:514–9. 10.1007/S12603-015-0599-4/FIGURES/2.

[25] Denier N, Clouston SAP, Richards M, Hofer SM. Retirement and cognition: A life course view. Adv Life Course Res 2017;31:11–21. 10.1016/j.alcr.2016.10.004.

[26] Teräs T, Rovio S, Pentti J, Head J, Kivimäki M, Stenholm S. Association of sleep with cognitive function during retirement transition: the Whitehall II study. Sleep 2023;46:1–9. 10.1093/SLEEP/ZSAC237.

[27] Okuda M, Noda A, Iwamoto K, Nakashima H, Takeda K, Miyata S, et al. Effects of long sleep time and irregular sleep-wake rhythm on cognitive function in older people. Sci Rep 2021;11. 10.1038/S41598-021-85817-Y.

[28] Swanson LM, Hood MM, Hall MH, Kravitz HM, Matthews KA, Joffe H, et al. Associations between sleep and cognitive performance in a racially/ethnically diverse cohort: the Study of Women’s Health Across the Nation. Sleep 2021;44. 10.1093/SLEEP/ZSAA182.

[29] Troxel WM, Haas A, Dubowitz T, Ghosh-Dastidar B, Butters MA, Gary-Webb TL, et al. Sleep Disturbances, Changes in Sleep, and Cognitive Function in Low-Income African Americans. Journal of Alzheimer’s Disease 2022;87:1591–601. 10.3233/JAD-215530.

[30] Teräs T, Myllyntausta S, Salminen M, Viikari L, Pahkala K, Muranen O, et al. The association of previous night’s sleep duration with cognitive function among older adults: a pooled analysis of three Finnish cohorts. Eur J Ageing 2023;20. 10.1007/S10433-023-00779-6.

[31] Stenholm S, Suorsa K, Leskinen T, Myllyntausta S, Pulakka A, Pentti J, et al. Finnish Retirement and Aging Study: a prospective cohort study. BMJ Open 2023;13. 10.1136/BMJOPEN-2023-076976.

[32] Teräs T, Rovio S, Spira AP, Myllyntausta S, Pulakka A, Vahtera J, et al. Associations of accelerometer-based sleep duration and self-reported sleep difficulties with cognitive function in late mid-life: the Finnish Retirement and Aging Study. Sleep Med 2020;68:42–9. S1389-9457(19)30318-1 [pii].

[33] Jenkins CD, Stanton BA, Niemcryk SJ, Rose RM. A scale for the estimation of sleep problems in clinical research. J Clin Epidemiol 1988;41:313–21. 0895-4356(88)90138-2 [pii].

[34] American Psychiatric Association. Diagnostic and Statistical Manual of Mental Disorders. Diagnostic and Statistical Manual of Mental Disorders 2013. 10.1176/APPI.BOOKS.9780890425596.

[35] BECK AT, WARD CH, MENDELSON M, MOCK J, ERBAUGH J. An inventory for measuring depression. Arch Gen Psychiatry 1961;4:561–71.

[36] Karasek RA. Job Demands, Job Decision Latitude, and Mental Strain: Implications for Job Redesign. Adm Sci Q 1979;24:285–308. 10.2307/2392498.

[37] Fransson EI, Nyberg ST, Heikkila K, Alfredsson L, Bacquer de D, Batty GD, et al. Comparison of alternative versions of the job demand-control scales in 17 European cohort studies: the IPD-Work consortium. BMC Public Health 2012;12:62. 10.1186/1471-2458-12-62 [doi].

[38] Health. FI of OH and FM of SA and. Riskikulutuksen varhainen tunnistaminen ja mini- interventio -hoitosuosituksen yhteenveto 2006.

[39] Zeger SL, Liang KY. Longitudinal data analysis for discrete and continuous outcomes. Biometrics 1986;42:121–30.

[40] Diggle P. The analysis of longitudinal data. Oxford: Oxford University Press; 2002.

[41] Vahtera J, Westerlund H, Hall M, Sjosten N, Kivimaki M, SalO P, et al. Effect of retirement on sleep disturbances: the GAZEL prospective cohort study. Sleep 2009;32:1459–66. 10.1093/sleep/32.11.1459 [doi].

[42] Roberts BA, Fuhrer R, Marmot M, Richards M. Does retirement influence cognitive performance? The Whitehall II Study. J Epidemiol Community Health (1978) 2011;65:958–63. 10.1136/jech.2010.111849 [doi].

[43] Zhang Q, Wu Y, Liu E. Longitudinal associations between sleep duration and cognitive function in the elderly population in China: A 10-year follow-up study from 2005 to 2014. Int J Geriatr Psychiatry 2021;36:1878–90. 10.1002/GPS.5615.

[44] Cacciamani F, Salvadori N, Eusebi P, Lisetti V, Luchetti E, Calabresi P, et al. Evidence of practice effect in CANTAB spatial working memory test in a cohort of patients with mild cognitive impairment. 2017;25:237–48. 10.1080/23279095.2017.1286346.

[45] Skirrow C, Cashdollar N, Granger K, Jennings S, Baker E, Barnett J, et al. Test-retest reliability on the Cambridge Neuropsychological Test Automated Battery: Comment on Karlsen et al. (2020). Appl Neuropsychol Adult 2022;29:889–92. 10.1080/23279095.2020.1860987.

