## Supplemental Figure 1 for "Concurrent changes in sleep and cognitive function during retirement transition: the Finnish Retirement and Aging Study"

**Supplemental Figure 1** Mean level and their 95% confidence intervals in each cognitive domain before and after retirement. Adjusted for age, sex, occupational position, depression, job strain, and alcohol consumption. The p-values that are shown are for the change in cognitive function during the retirement transition (wave -1 to 1).

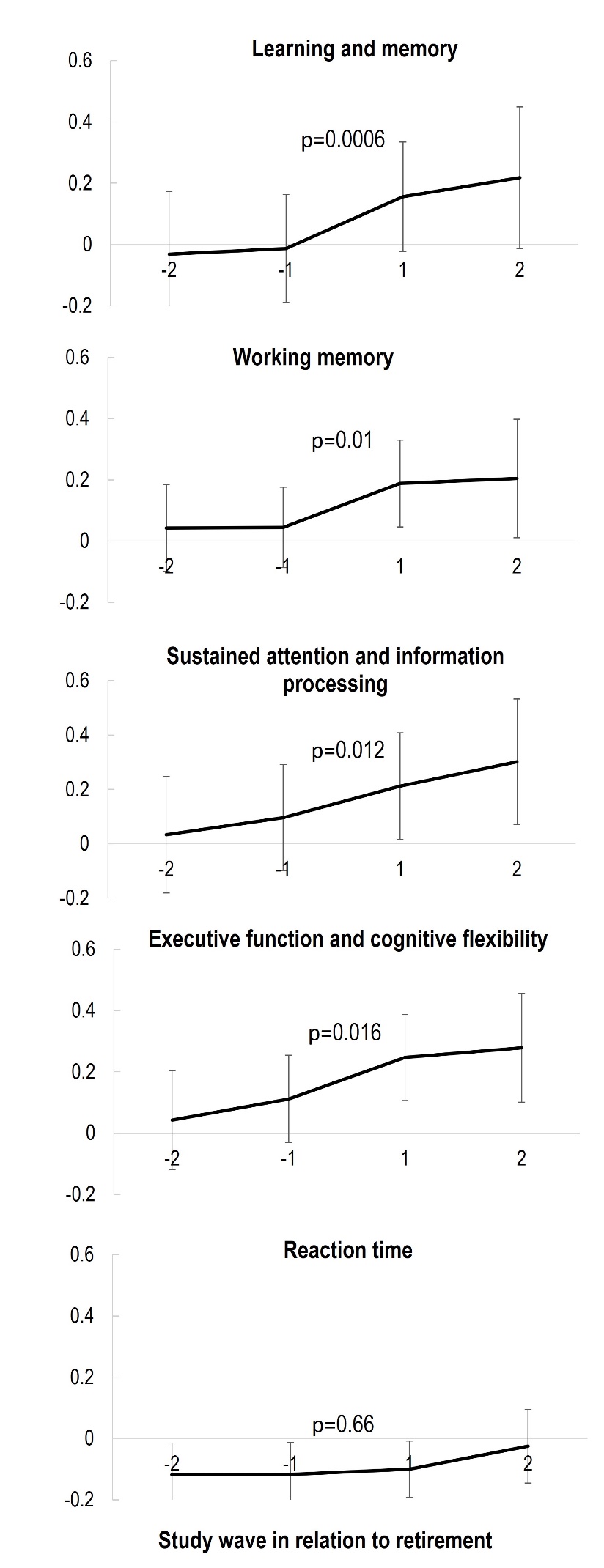
